# Surgery-induced reduction in inflammation relates to improved neural inhibitory control in obesity

**DOI:** 10.1101/2025.02.21.25322626

**Authors:** Lisa-Katrin Kaufmann, Emma Custers, Debby Vreeken, Jessica Snabel, Martine C. Morrison, Robert Kleemann, Maximilian Wiesmann, Eric J. Hazebroek, Esther Aarts, Amanda J. Kiliaan

## Abstract

Obesity is associated with impaired inhibitory control and low-grade systemic inflammation. Systemic inflammation adversely affects neurocognitive performance. Here, we investigate the effects of metabolic bariatric surgery on systemic inflammation and its influence on neural mechanisms underlying inhibitory control. In a sample of 47 individuals with obesity, we assessed inhibitory control processing pre- and 2 years post-bariatric surgery by probing neural activation and connectivity during an fMRI Stroop task. We investigated whether surgery-induced changes in plasma markers of systemic inflammation were related to changes in altered neural responses. Data were collected as part of the BARICO (Bariatric surgery Rijnstate and Radboudumc neuroimaging and Cognition in Obesity) study. Longitudinal analyses revealed decreased Stroop-related activation in the caudate nucleus and the left insula following surgery. These activation changes were accompanied by inflammation-related changes in functional coupling with medial superior frontal regions. Specifically, greater post-surgery decreases in leptin (pro-inflammatory) were associated with decreased connectivity between the anterior insula and the medial superior frontal regions, while increases in macrophage migration inhibitory factor (MIF, potentially neuroprotective) were linked to enhanced connectivity between the caudate nucleus and the medial superior frontal gyrus. Importantly, improved functional coupling between the caudate nucleus and the medial superior frontal gyrus was predictive of better task performance. Our findings suggest that surgery-induced reductions in systemic inflammation may improve inhibitory control in individuals with obesity by promoting neural changes in inflammation-sensitive brain regions and their functional interactions.

*This protocol was prospectively registered with the Dutch Trial Register Onderzoekmetmensen*.*nl, with trial number NTR29050*.

## 1 Introduction

Obesity is an increasingly prevalent condition that is associated with both significant deficits in inhibitory control and low-grade systemic inflammation (Chen et al., 2021; World Health Organization, 2024; Yang et al., 2018). To mitigate the serious health consequences of obesity, metabolic bariatric surgery can be used to induce weight loss (Colquitt et al., 2014). However, very little is known about how the surgery-related decrease of systemic inflammation impacts neurocognitive functioning.

Research has linked obesity to impairments in inhibitory control (Lavagnino et al., 2016; Restivo et al., 2017; Yang et al., 2018). Inhibitory control, the cognitive ability to suppress automatic responses or inappropriate behavior, can be assessed with response conflict tasks (Bari & Robbins, 2013). This ability is essential for goal-directed behavior, and deficits in inhibitory control are thought to contribute to both the development and maintenance of obesity (Nederkoorn et al., 2006; Stice & Burger, 2019; Yokum & Stice, 2023). Moreover, inhibitory control has been shown to predict treatment response and long-term outcomes in individuals with obesity (Stinson et al., 2018; Xu et al., 2017). At the neural level, fronto-parietal circuits and the insula have been proposed to play a central role in inhibitory control (Hung et al., 2018; Zhang et al., 2017). Emerging neuroimaging data suggests that alterations in neural activation in obesity may contribute to the observed behavioral deficits in inhibitory control, but findings are inconsistent and may be influenced by small sample sizes. Preliminary studies report reduced activation in regions including the supplementary motor area (SMA) and the insula during inhibitory control tasks like the stop signal task (Hendrick et al., 2012), and link higher body mass index (BMI) with lower insula activation (Filbey & Yezhuvath, 2017). Conversely, other studies report increased activation in these regions. For example, a small-scale study using a Stroop task observed increased activation in both the SMA and the insula in individuals with obesity (Balodis et al., 2013), while a Go/No-Go task study reports increased activation in the left insula and bilateral putamen among individuals with obesity compared to controls (Hsu et al., 2017). Taken together, these studies suggest inhibitory control-related deficits in several brain regions in individuals with obesity. However, findings remain inconclusive, and it is still unclear whether weight-loss treatment can improve these neurocognitive deficits.

These brain alterations are thought to be partly the effect of low-grade systemic inflammation associated with obesity (Li et al., 2023). Obesity is linked to low-grade systemic inflammation, evidenced by elevated levels of peripheral inflammation markers (Graßmann et al., 2017). Adipocyte expansion associated with excess body weight leads to increased leptin secretion and activation of inflammatory pathways, resulting in the upregulated expression of pro-inflammatory cytokines, including interleukin (IL)-6 (Park et al., 2010), C-C-motif chemokine ligand (CCL)3, and macrophage migration inhibitory factor (MIF) (Huber et al., 2008; Morrison & Kleemann, 2015; Vázquez-Moreno et al., 2020). This also induces increased C-reactive protein (CRP) production (Heredia et al., 2012). Such inflammatory processes in obesity have been linked to increased permeability of the blood-brain barrier and subsequent neuroinflammation (Guillemot-Legris & Muccioli, 2017). Importantly, mounting evidence suggests that systemic inflammation is linked to poorer neurocognitive performance (Chen et al., 2021; Shi et al., 2022) and influences brain activation in key areas responsible for inhibitory control, such as the prefrontal cortex and the striatum (Kraynak et al., 2018). Understanding how inflammation mediates the impairment of inhibitory control could provide valuable insights into the neurocognitive effects of obesity.

Effective treatment for obesity aims to achieve long-term weight loss. Metabolic bariatric surgery is a way to induce rapid and sustained weight loss by reducing stomach size and limiting food intake and nutrient absorption (Arterburn et al., 2020; Colquitt et al., 2014). Surgery-induced weight loss can be beneficial for cognitive function and significantly reduces inflammation (Handley et al., 2016; Thiara et al., 2017; Vreeken et al., 2023). Moreover, decreases in inflammation have been found to predict improvements in cognitive function (Vreeken et al., 2023). However, the neural mechanisms through which surgery-induced decreases in systemic inflammation impact neurocognitive functioning remain unclear.

Here, we employed a longitudinal design to identify long-term changes in both systemic inflammation and brain parameters in individuals with obesity. We examined inhibitory control processing before and 2-years after metabolic bariatric surgery, by probing neural responses during the Stroop task (Stroop, 1935), a well-established measure of inhibitory control (Cieslik et al., 2015). First, we investigated whether surgery-induced changes in key markers of systemic inflammation were linked to changes in neural activation during inhibitory control processing, focusing on brain regions sensitive to inflammation (see Kaufmann et al., 2024). Second, we investigated associated changes in functional connectivity during inhibitory control processing, to assess alterations of neural circuits required for task performance. Third, we investigated whether inflammation-related changes in neural responses were linked to changes in behavior. The findings provide insights into the relationship between long-term changes in systemic inflammation and brain parameters, elucidating the mechanisms underlying cognitive improvement following weight loss.

## 2 Methods and Materials

### 2.1 Participants

Seventy-five women and men (aged 35-55) with severe obesity, i.e., BMI > 40kg/m^2^ or > 35kg/m^2^ with comorbidities (Fried et al., 2008), were enrolled in the study prior to metabolic bariatric surgery. Participants were recruited as part of the larger study protocol of the *BARICO* (Bariatric surgery Rijnstate and Radboudumc neuroimaging and Cognition in Obesity) project (Vreeken et al., 2019). 62 participants completed the pre-surgery assessment and the 2-year follow-up, twelve of which had to be excluded due to insufficient accuracy on the Stroop task (< 50%). Three additional participants were excluded due to severely elevated inflammation levels (see clinical measures). A total of 47 individuals (seven men, 15%) were included in the analyses (**Figure 1A)**. Details are provided in **Supplemental Methods**.

**Figure 1.**
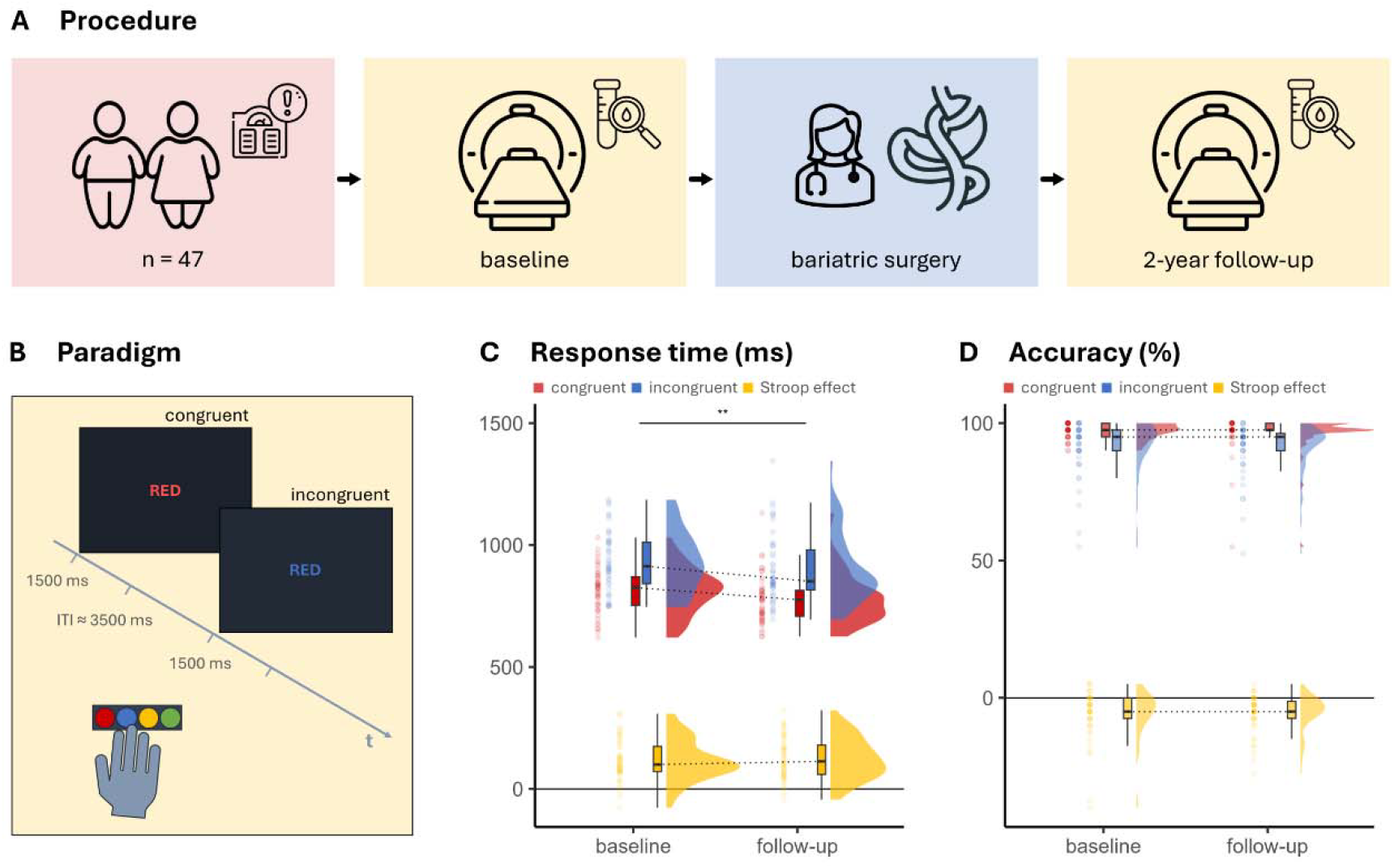
Overview of study design and surgery-related changes in performance. **A Procedure.** Patients qualifying for metabolic bariatric surgery were scanned before and 2 years after surgery. Blood samples were collected at each time point to assess inflammation markers. **B Color-word Stroop task paradigm.** In the congruent condition, the color of the ink and the color word matched. In the incongruent condition, there was a mismatch between the color of the ink and the color word. The duration of each stimulus lasted 1500ms and the inter-trial interval (ITI) ranged from 2000 to 4000ms. Participants were asked to respond to the ink color – instead of the meaning of the word – as fast and accurately as possible by pressing color-coded keys with the right hand. The order of words was pseudorandomized and counterbalanced across participants. **C Response time results.** The Stroop effect was calculated as the difference in mean response time of correct trials between the incongruent and the congruent condition. Response time across both conditions decreased statistically significantly after surgery, indicating participants were responding faster at follow-up. ** *p <* 0.01. **D Accuracy results.** The Stroop effect was calculated as the difference in mean accuracy of trials between the incongruent and the congruent condition. There was no evidence for a change in accuracy between sessions. Boxplots show the median as horizontal marker, the edges of the box represent the 25th and 75th percentiles, and the whiskers span 1.5 interquartile ranges.

This sample overlaps with previous studies of the BARICO project (Custers et al., 2023, 2024; Kaufmann et al., 2024; Vreeken et al., 2022, 2023). All participants provided written informed consent prior to participation. The study protocol complied with the Declaration of Helsinki and the ICH Harmonised Tripartite Guideline for Good Clinical Practice, was approved by the local ethics review board, the medical review ethics committee (METC Oost-Nederland NL63493.091.17), and was registered in the Dutch Trial Register (protocol number: NTR29050).

### 2.2 Clinical measures

Current height and weight were used to calculate participants’ BMI as a measure of obesity. Further anthropometric measures included waist circumference, as a measure of abdominal adiposity, and percentage of total body weight loss (TBWL%). Depressive symptoms were assessed with the Dutch version of the Beck Depression Inventory (BDI) (Beck & Steer, 1993). Blood samples were collected at each time point to assess inflammation markers, including CRP, leptin, IL-6, IL-8, CCL3, and MIF levels. Inflammatory markers were chosen based on previous work of our group, which explored associations with obesity and depression (Kaufmann et al., 2024). To ensure that elevated inflammation levels at baseline were due to chronic rather than acute inflammation, the latter being indicated by severely elevated values, participants with inflammation levels exceeding 3 standard deviations above the mean were excluded. Further details are provided in **Supplemental Methods**. To examine changes in clinical measures, paired Welch’s t-tests were calculated between baseline and follow-up measures. Four of the six inflammation markers showed significant changes at follow-up and were considered for further analyses. Associations between changes in clinical measures were calculated as partial Spearman’s rank correlations, controlling for age and sex (assigned at birth), using R (RRID:SCR_001905, version 4.4).

### 2.3 Stroop task

Inhibitory control (i.e., response conflict processing) was assessed at both time points using a Dutch version of the color-word Stroop paradigm during fMRI. Participants were asked to indicate the color of four color words, presented either in the same color as the word (*congruent condition*, e.g. ‘red’ in red ink) or in a different color (*incongruent condition*, e.g. ‘red’ in blue ink, **Figure 1B)**, by pressing a corresponding button and ignoring the meaning of the word. Possible colors were red, blue, yellow, and green. To familiarize themselves with the task and the color-button contingency, participants first performed 10 practice trials with feedback. Subsequently, the task consisted of a total of 80 trials and lasted approximately 10 minutes. The task was delivered using Presentation (RRID:SCR_002521). All stimuli and the task code are available at the Open Science Framework (https://dx.doi.org/10.17605/osf.io/c5dvh).

Mean response time (RT) of correct trials and response accuracy were analyzed as measures of task performance (**Figure 1C,D)**. Response times were trimmed before analysis to eliminate anticipation or late responses, considering response times between 200ms and 3 standard deviations of the participant-and-condition-specific mean response time as valid trials (excluding < 0.01% of all data). Paired Welch’s t-tests were calculated to compare baseline and follow-up measures.

### 2.4 Statistical analyses

The statistical analyses had three aims. First, we investigated brain activation changes within distinct inflammation-sensitive regions relevant for inhibitory control processing using ROI-wise linear mixed-models. Second, we investigated inflammation-specific changes of functional connectivity within these inflammation-sensitive regions. Third, we investigated whether inflammation-related changes in neural responses were linked to changes in behavior. Scripts for the main analyses are available at the Open Science Framework (https://doi.org/10.17605/osf.io/zxtks).

### 2.5 Analysis of fMRI activation

Functional images were subjected to preprocessing procedures, including realignment, slice timing correction, spatial normalization, and smoothing. We generated a single contrast image per participant for the following comparisons: the main task effect [incongruent > congruent] at baseline [incongruent_baseline_ - congruent_baseline_] and follow-up [incongruent_follow-up_ - congruent_follow-up_], and the change of activation over time [follow-up > baseline] using the contrast [(incongruent_follow-up_ - congruent_follow-up_) - (incongruent_baseline_ - congruent_baseline_)]. Second-level analyses comprised one-sample t-tests for the contrasts [incongruent > congruent] at both baseline and follow-up, and for the [follow-up > baseline] contrast, with age and sex as covariates of no interest. Statistically significant activation clusters were identified using a cluster-defining threshold of p < 0.001 and a minimum spatial extent of 10 voxels, controlling the family-wise error (FWE) rate at α ≤ 0.05 across inflammation-sensitive brain regions (Kraynak et al., 2018). Details are provided in **Supplemental Methods**.

Signal level within the five clusters showing the strongest activation differences at baseline were calculated by extracting the mean beta values from participants’ first-level contrast maps for the [incongruent > congruent] comparisons at baseline and follow-up. These values were then used for linear mixed-effects analyses. The mixed models included brain activation as dependent variable, with time, age, and sex as fixed effects, and a random intercept for each participant. The significance level for the mixed-effects analyses was set to ⍰α ≤ ⍰0.05, FDR-corrected for multiple regions of interest (Benjamini & Hochberg, 1995).

Results were labeled according to the Automated Anatomical Labeling atlas 3v1 (Rolls et al., 2020) using AtlasReader (Notter et al., 2019). Results were visualized as dual-coded images (Allen et al., 2012) using the *nanslice p*ackage (https://github.com/spinicist/nanslice). The unthresholded T-maps representing the neural activation for the contrast [incongruent > congruent] at baseline and follow-up are available on NeuroVault (RRID:SCR_003806; https://identifiers.org/neurovault.image:887947).

### 2.6 Analysis of fMRI connectivity

We examined the functional connectivity of response conflict sensitive regions during inhibitory control using generalized psychophysiological interaction (gPPI) analyses (RRID: SCR_009489; McLaren et al., 2012), with the five clusters showing the largest differences in activation between conditions at baseline serving as seed regions. Details are provided in **Supplemental Methods**. A second-level analysis used one-sample t-tests on the contrast images to probe connectivity changes during response conflict, including age and sex as covariates of no interest. Statistically significant clusters of task-modulated connectivity were obtained using a cluster-defining threshold of p < 0.001, controlling the family-wise-error rate at α ≤ 0.05 across the target space of inflammation-sensitive brain regions (Kraynak et al., 2018). To probe whether changes in inflammation markers were linked to changes in brain connectivity during response conflict, we included change scores of CRP, leptin, IL-6, and MIF as regressors in separate second-level GLMs. These analyses used a cluster-defining threshold of *p <* 0.001 and controlled the FWE rate at α ≤ 0.0125 across the target space and the four comparisons. Results were labelled according to the Automated Anatomical Labeling atlas 3v1 (Rolls et al., 2020). Unthresholded T-maps representing the inflammation-specific changes in task connectivity for the contrast [follow-up > baseline] are available on NeuroVault (RRID:SCR_003806; https://identifiers.org/neurovault.collection:17746).

### 2.7 Inflammation-behavior, inflammation-brain-, and brain-behavior analyses

To investigate associations between participants’ neural measures, task performance, and inflammation markers we used linear mixed-effect models. Model assumptions, including normality of residuals, homoscedasticity, and linearity, were thoroughly checked. In cases where these assumptions were violated, Spearman rank correlations were calculated using the residuals of the change scores, controlling for age and sex. This approach provided a robust alternative to examine the relationships between variables, while mitigating the impact of assumption violations.

For the inflammation-behavior analyses, we used linear mixed-effect models with the accuracy or response time Stroop effect as dependent variable. Inflammation markers (one at a time), time, age, and sex were included as fixed effects, with a random intercept for each participant. For the inflammation-brain and brain-behavior analyses, we calculated signal levels within brain activation and connectivity clusters that showed significant change between baseline and follow-up, by extracting the mean beta values from participants’ first-level contrast maps for the [incongruent > congruent] comparisons at both time points. We employed linear mixed-effect models with brain activation or connectivity as dependent variable. Inflammation or performance markers (one at a time), time, age, sex, and the interaction between inflammation/performance marker and time were fixed effects, with a random intercept for each participant.

All fixed effects parameters were z-standardized to facilitate interpretation and improve numerical stability. Linear mixed effects modeling was performed in R (RRID:SCR_001905, version 4.4), using the R packages *lme4 (*RRID:SCR_015654) and *lmerTest (*RRID:SCR_015656). The significance level for the mixed-effects analyses was set to ⍰α ≤ ⍰0.05, FDR-corrected for multiple regions of interest and inflammation markers (Benjamini & Hochberg, 1995).

## 3 Results

### 3.1 Demographic and anthropometric results

Descriptive statistics of the participants are summarized in **Table 1**. At baseline, 20 participants (43%) of the sample scored above the cut-off for mild to moderate depression (BDI score > 9). At the follow-up assessment, 3 participants (6.7%) remained above this threshold.

**Table 1.** Demographic and clinical characteristics.

| Characteristic | baseline, N = 47 | follow-up, N = 47 | p-value |
| --- | --- | --- | --- |
|  | Mean (SD), [Range] / n (%) | Mean (SD), [Range] / n (%) | paired t-test |
|  | Mean (SD), [Range] / n (%) | Mean (SD), [Range] / n (%) | paired t-test |
| Age | 45.4 (6.3, [36.2, 55.2]) | - |  |
| Sex assigned at birth (female / male) | 40 (85%) / 7 (15%) | - |  |
| Education level (low / middle / high)<sup>1</sup> | 5 (11%) / 20 (43%) / 22 (47%) | - |  |
| BMI (kg/m<sup>2</sup>) | 40.3 (2.6), [34.7, 47.8] | 26.1 (2.8), [19.0, 31.3] | <0.001 |
| Waist circumference (cm) | 121.0 (9.7), [106.0, 145.0]<sup>2</sup> | 91.3 (9.8), [70.0, 118.0]<sup>3</sup> | <0.001 |
| Total body weight loss (TBWL%) | - | 34.9 (6.8), [22.9, 51.3] |  |
| Depression (BDI) | 8.8 (5.0), [1.0, 22.0]<sup>3</sup> | 3.6 (4.0), [0.0, 21.0]<sup>4</sup> | <0.001 |
| **Medication use** |  |  |  |
| Oral antidiabetics | 5 (11%) | 2 (4.3%)<sup>3</sup> |  |
| Antihypertensives | 11 (23%) | 3 (6.4%)<sup>3</sup> |  |
| Insulin therapy | 3 (6.4%) | 1 (2.1%)<sup>3</sup> |  |

<sup>1</sup> According to the Verhage score (Verhage, 1964): low ≤ 4, middle = 5, high ≥ 6, based on the Dutch educational system, akin to the International Standard Classification of Education (UNESCO Institute for Statistics, 2012).
<sup>2</sup> n=40, <sup>3</sup> n=46, <sup>4</sup> n=45

### 3.2 Inflammation results

Before surgery, 32 participants (68%) had elevated CRP levels (> 3mg/l), a crude measure for systemic inflammation (Ishii et al., 2012). At the 2-year follow-up, only 3 participants (6.4%) still showed elevated CRP levels. Both leptin and IL-6 levels also decreased significantly after surgery, while MIF levels showed a significant increase. These four inflammation markers with significant changes at follow-up were considered for subsequent analyses examining relationships with behavior and brain changes. Levels of all inflammatory markers are summarized in **Table 2**. Decreased leptin levels were associated with decrease in BMI (*rho* = 0.47, *p <* 0.001, *q <* 0.004; 95%-CI [0.20, 0.67]) and TBWL% (*rho* = -0.49, *p* < 0.001, *q* < 0.002; 95%-CI [-0.69, -0.23]), while the other markers showed no such associations.

**Table 2.** Inflammation levels at baseline and follow-up.

| Characteristic | baseline, N = 47 | follow-up, N = 47 | p-value |
| --- | --- | --- | --- |
|  | Mean (SD), [Range] | Mean (SD), [Range] | paired t-test |
| CRP (ug/ml) | 6.08 (5.35), [0.49, 26.69] | 1.19 (1.46), [0.10, 6.88] | <0.001 |
|  | Mean (SD), [Range] | Mean (SD), [Range] | paired t-test |
| Leptin (ng/ml) | 64.38 (24.83), [17.16, 136.77] | 14.55 (10.54), [1.30, 57.09] | <0.001 |
| IL-6 (pg/ml) | 2.15 (1.11), [0.42, 5.36] | 0.63 (0.67), [0.02, 3.19] | <0.001 |
| IL-8 (pg/ml) | 8.40 (3.58), [2.87, 17.40] | 9.78 (4.61), [2.41, 25.06] | 0.055 |
| MIF (ng/ml) | 11.60 (8.20), [1.85, 39.61] | 21.92 (13.42), [4.94, 76.21] | <0.001 |
| CCL3 (pg/ml) | 18.44 (7.50), [0.00, 41.96] | 19.42 (8.61), [4.23, 62.24] | 0.4 |

### 3.3 Behavioral results

Comparing the Stroop task conditions, we observed a robust Stroop effect with a large effect size for response time (*baseline: Δ r*esponse time = 116.86 ± 84.05, t(46) = 9.53, *p <* 0.001, *d* =1.39; *follow-up: Δ r*esponse time = 124.59 ± 85.45, t(46) = 10.00, *p <* 0.001, *d* =1.46) and a moderate to large effect size for response accuracy (*baseline: Δ a*ccuracy = -5.96 ± 9.22, t(46) = -4.43, *p <* 0.001, *d* = -0.65; *follow-up: Δ a*ccuracy = -5.53 ± 6.84, t(46) = - 5.55, *p <* 0.001, *d* = -0.81). These results confirmed the presence of response automaticity at both sessions (**Figure 1C,D)**. Comparative analysis between the sessions revealed a statistically significant decrease in response time across conditions, indicating that participants were responding faster at follow-up (mean difference = -34.05 ± 85.78, t(46) = -2.721, *p <* 0.01, *d* = -0.40) (**Figure 1C)**. There was no evidence of change in the Stroop effect for response time or accuracy over time (all *p >* 0.50). However, there was a trend suggesting that individuals with a greater decrease in CRP also showed a greater improvement in the Stroop accuracy effect, indicating improved performance (*rho* = -0.32, 95%-CI [-0.56, -0.03], *p* = 0.014, *q* = 0.056). Changes in other inflammation markers were not associated with changes in performance (all *p >* 0.40).

### 3.4 Neuroimaging results

#### 3.4.1 Changes in brain activation during response conflict

*W*e identified inhibitory control processing areas as brain regions with greater activation for the [incongruent > congruent] GLM contrast within a mask of inflammation-sensitive brain regions, controlling for age and sex of participants (**Figure 2)**. At baseline, this contrast revealed prominent activation in brain regions associated with response conflict processing (Langner et al., 2018), with the five strongest located in the left SMA, the left middle frontal gyrus (MFG), the left anterior insula extending to the putamen and caudate, the right anterior insula, and the right caudate nucleus (**Figure 2B; s**ee **Table S2 f**or a summary of all clusters and coordinates).

**Figure 2.**
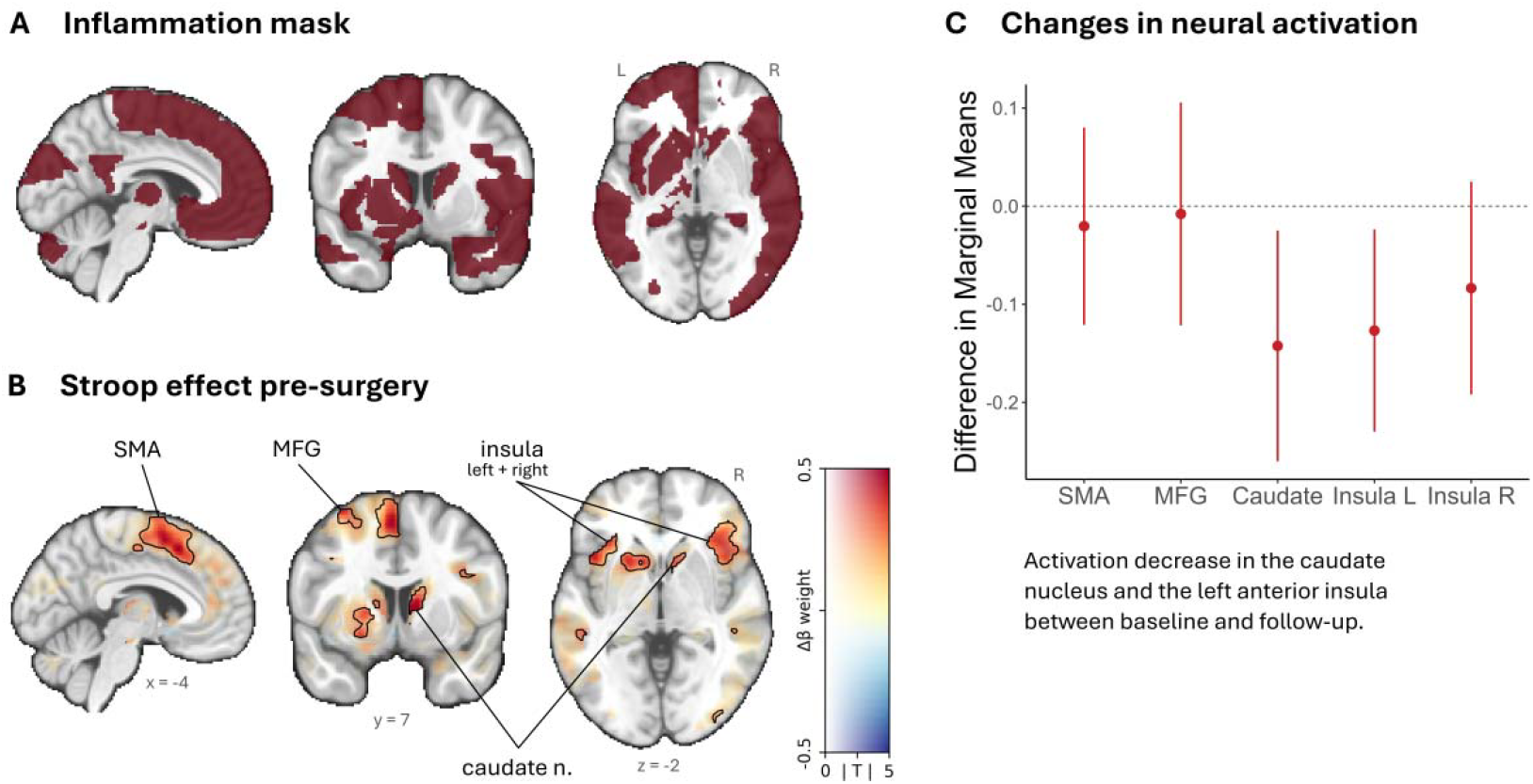
Neural activation during inhibitory control processing. **A** The Stroop effect was calculated within a mask of inflammation-sensitive brain regions (Kraynak et al., 2018), see Supplemental Methods for further details. **B** During inhibitory control processing at baseline, participants showed greater activation in several brain regions, including the left supplementary motor area (SMA), the left middle-frontal gyrus (MFG), the left and right insula, and the right caudate nucleus. Dual-coded contrast: color indicates the estimated regression coefficient (vertical axis of color bar) and transparency corresponds to the absolute t-statistic values (horizontal axis of color bar). Significant clusters (*p*FWE < 0.05) are contoured black to facilitate the interpretation. See **Table S2** for a full list and Montreal Neurological Institute (MNI) coordinates of significant clusters. **C** Neural activation within the caudate nucleus and the left insula showed a significant decrease across time (follow-up > baseline).

Based on these results, we investigated changes in task-related brain activation of the left SMA, the left MFG, the left and right insula, and the right caudate nucleus between baseline and the 2-year follow-up. Linear mixed-effects analyses revealed a significant decrease in activation over time in the caudate nucleus (beta = -0.14, 95% CI [-0.26, -0.03], *t(*88) = -2.44, *p* = 0.017, *q* = 0.04) and left insula (beta = -0.13, 95% CI [-0.23, -0.03], *t(*88) = -2.48, *p* = 0.015, *q* = 0.04) (**Figure 2C)**.

Next, we investigated the association between brain activation and inflammation measures (CRP, leptin, IL-6, and MIF). Linear mixed-effect models showed no statistically significant effects (all *p >* 0.06) (**Table S3)**. Further linear mixed-effects analyses examining associations between activation and performance measures showed no evidence for an association between brain activation and the Stroop effects (**Table S4)**.

#### 3.4.2 Changes in task connectivity during response conflict

*T*o investigate changes in functional connectivity, we conducted gPPI analyses using the five clusters showing the strongest peak activation at baseline in the [incongruent>congruent] GLM contrast as seed regions (**Figure 2B+3B)** within the mask of inflammation-sensitive brain regions (**Figure 2A)**. There was no evidence for general connectivity changes within these regions.

We next examined specific inflammation-related connectivity changes for CRP, leptin, IL-6, and MIF, to assess the influence of surgery-induced changes in individual inflammation levels (**Figure 3A, Table S5)**.

**Figure 3.**
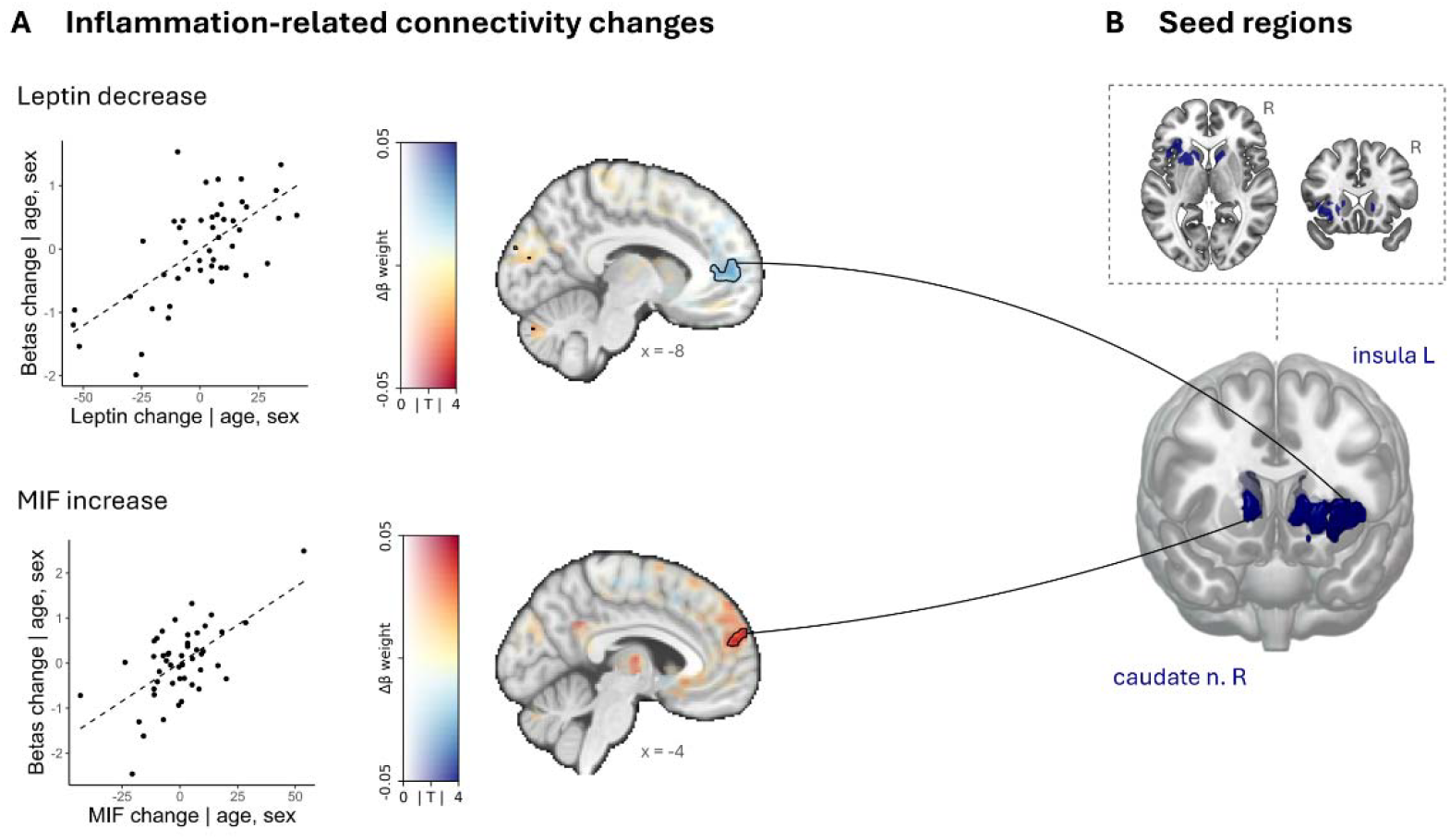
Change in conflict-related functional connectivity between baseline and the 2-year follow-up. **A** Inflammation-related connectivity analyses showed decreased leptin values being related to decreased connectivity between the left insula and the left medial superior frontal gyrus. Increased MIF levels were associated with increased connectivity between the caudate nucleus and the medial part of the left superior frontal gyrus. Dual-coded contrast: color indicates the estimated regression coefficient (vertical axis of color bar) and transparency corresponds to the absolute t-statistic values (horizontal axis of color bar). Significant clusters (*p*FWE < 0.05) are contoured black to facilitate the interpretation. **B** Seed regions. See **Table S5** for a full list and Montreal Neurological Institute (MNI) coordinates of significant clusters.

Participants with greater decrease in leptin showed a greater decrease in connectivity between the left insula and the medial part of the left superior frontal gyrus (SFGmed) (*k* = 138, *p*FWE < 0.001). Furthermore, a greater increase in MIF was linked to a greater increase in connectivity between the right caudate nucleus and the left SFGmed (*k* = 121, *p*FWE < 0.004).

Notably, subsequent mixed-model analyses revealed that higher MIF-related connectivity was associated with a smaller (i.e. less negative) accuracy Stroop effect (beta = 0.37, SE = 0.15, t(86) = 2.42, *p* = 0.018, *q* = 0. 036), indicating better performance in the incongruent condition. This effect was driven by baseline, as indicated by a significant interaction effect (β = -0.70, SE = 0.26, t(86) = -2.73, p = 0.008, *q* = 0. 015) (**Table S6)**.

## 4 Discussion

Controlling automatic or prepotent responses is an essential skill for navigating situations with competing demands. Individuals with obesity often exhibit deficits in overriding automatic response tendencies, known as inhibitory control (Lavagnino et al., 2016; Restivo et al., 2017; Yang et al., 2018). Low-grade systemic inflammation, which is prevalent in obesity, has been associated with increased deficits (Chen et al., 2021; Shi et al., 2022). Little is known regarding the influence of metabolic bariatric surgery-induced decreases in systemic inflammation on the neural processing of inhibitory control in individuals with obesity. Results from the current study revealed significant surgery-induced changes in inflammation markers, alongside decreased brain activation in inflammation-sensitive regions (Kraynak et al., 2018), specifically the caudate nucleus and the left insula. These activation changes were accompanied by inflammation-related changes in functional coupling with medial superior frontal regions, brain areas associated with inhibition, decision-making, and self-control (Euston et al., 2012). Crucially, functional coupling during inhibitory control processing between the caudate nucleus and the SFGmed predicted better task performance, with increased crosstalk in individuals with better task accuracy. Our findings provide evidence that surgery-induced decreases of systemic inflammation may improve inhibitory control in individuals with obesity by altering functional coupling in fronto-striatal regions.

### 4.1 Inflammation-sensitive regions show decreased activation after surgery

Following surgery, we observed a decrease in brain activation within inflammation-sensitive regions during inhibitory control processing following surgery, along with reduced inflammation measures. Specifically, activation in the right caudate nucleus and left anterior insula was significantly lower at follow-up than at baseline. This decrease in activation may reflect a normalization process, as previous studies have shown that individuals with obesity exhibit heightened activation in regions such as the insula, caudate nucleus, and putamen during response inhibition tasks compared to non-obese controls (Balodis et al., 2013; Hsu et al., 2017). Although the relatively long two-year interval between assessments makes learning effects on these findings less likely, we cannot fully rule out general test-retest effects due to the lack of a control group. Nonetheless, evidence from a case-controlled study in adolescents also shows improved executive function and post-surgical reductions in brain activation during an N-back task in the anterior insula (Pearce et al., 2017). Moreover, meta-analyses of behavioral data support improvements in executive function following weight-loss interventions, including metabolic bariatric surgery (Siervo et al., 2011; Thiara et al., 2017), though such performance improvements were not found currently. Taken together, these results suggest that post-surgery weight loss may alleviate hyperactivation in brain regions involved in inhibitory control. The observed decrease in activation post-surgery may indicate more efficient processing of response conflicts, requiring less neural activity for the same level of performance.

### 4.2 Inflammation-specific connectivity changes

Functional circuit analyses revealed inflammation-specific connectivity changes between control-processing regions and the SFGmed. Notably, regions with a significant post-surgery decrease in activation also showed inflammation-specific changes in connectivity. Specifically, a greater decrease in leptin levels was associated with decreased connectivity between the SFGmed and the left insula, while a greater increase in MIF was linked to increased connectivity between the SFGmed and the right caudate nucleus. The SFGmed is implicated in a wide range of processes and is thought to function as a central hub, integrating and coordinating adaptive behavior (Roy et al., 2012). It plays a critical role in decision-making processes, such as response conflict monitoring (Alexander & Brown, 2011; Botvinick et al., 2004), and supports the stable maintenance of task control (Dosenbach et al., 2007, 2008).

The **leptin-associated decrease** in functional connectivity between the insula and the SFGmed aligns with findings from leptin substitution therapy in lipodystrophy, where leptin was positively associated with intrinsic connectivity in regions including the insula and the medial prefrontal cortex (Schlögl et al., 2016). Leptin exerts a pro-inflammatory effect on the body (Abella et al., 2017) and is closely linked with adipose tissue mass, with levels known to decrease following weight loss (Maffei et al., 1995). Peripheral inflammation markers such as leptin can modulate fronto-striatal circuits both by directly acting on receptors in the striatum and by indirect effects via the ventral tegmental area (Janssen et al., 2019). The anterior insula, known to play a role in attentional control (Nelson et al., 2010), has a close functional and anatomical relationship with medial prefrontal regions (Dosenbach et al., 2007; Nelson et al., 2010). Given the functional role of SFGmed-insula connectivity in stable task control (Dosenbach et al., 2007; Nelson et al., 2010) and the link between improved leptin levels and better post-surgery cognitive function (Alosco et al., 2015; Nozari et al., 2023), the observed leptin-associated decrease in functional coupling between these regions suggests that surgery-induced reductions in leptin may contribute to a shift towards more adaptive neural processing patterns.

The **MIF-associated increase** in functional connectivity between the caudate nucleus and the SFGmed is consistent with reports on obesity, where higher BMI has been associated with lower intrinsic connectivity between the right caudate nucleus and the SFGmed/anterior cingulate cortex (Zhao et al., 2021). The SFGmed, with its prominent unilateral projections to the caudate nucleus (Euston et al., 2012), demonstrates hyperconnectivity with the caudate in conditions characterized by excessive control behaviors, such as obsessive-compulsive disorders (Apergis-Schoute et al., 2018). As obesity is associated with deficits in cognitive control, the observed increase in connectivity may reflect improvements in neurocognitive functioning. In fact, the MIF-associated increase in connectivity was linked to improved task performance, evidenced by higher accuracy during the incongruent condition of the Stroop task. While MIF is typically recognized for its pro-inflammatory role in obesity progression (Morrison & Kleemann, 2015), it also exerts protective effects against metabolic stress through mechanisms influenced by specific cellular contexts (Leyton-Jaimes et al., 2018; Morrison & Kleemann, 2015). The post-surgical increase in MIF observed in this study aligns with prior reports of weight-loss-related MIF increases in obesity (van Dielen et al., 2004) (but see also Kleemann and Bucala (2010)). Given the link between MIF-associated connectivity changes and performance improvement, it appears that the protective effects of MIF may be more prominent in this cohort. These results indicate that post-surgical increases in MIF may facilitate improved neurocognitive performance.

The observed performance improvements in response time in this study align with meta-analytic findings showing improved cognitive function, including executive function as measured by the Stroop task, 12 months post-surgery (Tao et al., 2024), as well as reports of sustained executive function improvements three to four years after surgery (Alosco et al., 2014). Together, these findings suggest that weight-loss-related reductions in inflammation may promote more efficient inhibitory control processing by reshaping functional coupling of inflammation-sensitive brain regions. Obesity-induced systemic inflammation appears to have a substantial but reversible impact on brain function during inhibitory control. Overall, these findings indicate that sustained weight loss may help reverse cognitive impairments linked to elevated BMI (Bocarsly et al., 2015; Hendrick et al., 2012; Vainik et al., 2013), highlighting the potential cognitive benefits of reducing obesity-related systemic inflammation.

### 4.3 Limitations

The lack of a control group in the present study limits our ability to draw definitive causal conclusions about the effects of metabolic bariatric surgery on systemic inflammation and neural connectivity. In addition, the sample of this study comprised more women than men, precluding any subgroup analyses. Future studies would benefit from a more balanced sample, allowing the investigation of potential sex differences (Braga Tibaes et al., 2024; Lasselin et al., 2018; ter Horst et al., 2020).

## 4.4 Conclusion

The use of a longitudinal design enabled us to identify long-term changes in both systemic inflammation and brain parameters. Our findings suggest that surgery-induced decreases of systemic inflammation may improve inhibitory control in individuals with obesity through neural mechanisms involving inflammation-sensitive brain regions and their crosstalk. This underscores the potential cognitive benefits of weight loss interventions in managing obesity and highlights the importance of addressing systemic inflammation in therapeutic strategies.

## Data Availability

All data from this study are available upon request from the last authors. All stimuli, the stimulus presentation scripts, and scripts for the main analyses are available at https://doi.org/10.17605/osf.io/h846n.

## Data and code availability

Data from this study are available upon request from the last authors. All stimuli, the stimulus presentation scripts, and scripts for the main analyses are available at https://doi.org/10.17605/osf.io/h846n.

## CRediT author statement

LKK: conceptualization, formal analysis, visualization, writing – original draft, review and editing. EC: investigation, data curation, writing – review and editing. DV: investigation, data curation. JS: methodology (biomarker assay optimization, matrix effects), investigation (biomarker measurements and analysis). MM: methodology (biomarker assay development), supervision (biomarker analysis). RK: conceptualization (biomarker assay development), resources, supervision, writing – review and editing. MW: supervision, writing – review and editing. EH: supervision, writing – review and editing. EA: conceptualization, funding acquisition, resources, supervision, writing – review and editing. AK: conceptualization, funding acquisition, resources, supervision, writing – review and editing. All authors contributed to the article and approved the submitted version.

## Acknowledgments

We thank all participants who participated in our study and A. Hofboer for her assistance with participant recruitment and data collection. Further, we are grateful to W. van Duyvenvoorde, S. Drift-Dongen, E. Gart, and F. Seidel for technical assistance with the blood analyses.

## Ethical Standards

The authors assert that all procedures contributing to this work comply with the ethical standards of the relevant national committees on human experimentation, the METC Oost-Nederland (NL63493.091.17), with the Helsinki Declaration of 1975, as revised in 2008, and the ICH Harmonised Tripartite Guideline for Good Clinical Practice. The study was registered in the Dutch Trial Register (protocol number: NTR29050).

