## Supplement for "Surgery-induced reduction in inflammation relates to improved neural inhibitory control in obesity"

#### *- Supplemental Information -*

#### Supplemental Methods

##### Participants

All participants were right-handed and had sufficient command of Dutch. Exclusion criteria comprised previous or current neurological or psychiatric illness; pregnancy; treatment with antibiotics, probiotics or prebiotics 3 months before or during the study (excluding preoperative prophylaxis); color blindness; and standard exclusion criteria for the MRI, i.e., claustrophobia; epilepsy; pacemakers or defibrillators; nerve stimulators; intracranial clips; infraorbital or intraocular metallic fragments; cochlear implants; ferromagnetic implants; and circumference above the MRI space capacity (Vreeken et al., 2019).

Participants underwent Roux-en-Y gastric bypass surgery, where a small gastric pouch is connected to the small intestine, bypassing the stomach, duodenum, and the proximal part of the jejunum. This restrictive procedure aims to facilitate long-term weight loss (Arterburn et al., 2020; Peterli et al., 2018).

##### Clinical measures

*Anthropometric measures*

Anthropometric measurements included Body Mass Index (BMI), waist circumference, and percentage total body weight loss (TBWL%), calculated as $\frac{(\text{weight}_{\text{baseline}} - \text{weight}_{2\text{yr}})}{\text{weight}_{\text{baseline}}} \text{x } 100$. Body weight was measured with participants wearing clothes and shoes, with jackets removed. To correct for the clothing and shoes, 1 kg was subtracted from the measured weight. Height was measured with shoes removed.

*Psychometric measures*

Depressive symptoms were assessed with the Dutch version of the Beck Depression Inventory (BDI) version IA (Beck & Steer, 1993), consisting of 21 questions about the participant's feelings over the past week. Responses were scored on a scale from 0 to 3 and summed to calculate the total score, ranging from 0 to 63. Higher scores reflect a higher level of depression symptoms and negative affect (Watson & Clark, 1984). The following cut-off scores can be applied: 0-9 for minimal depression, 10-18 for mild depression, 19-29 for moderate depression, and 30 or higher for severe depression (Beck et al., 1988). Cronbach's alpha for the present sample was α = 0. 76 (bootstrap 95%-CI [0.65, 0.83]) at baseline and α = 0. 80 (bootstrap 95%-CI [0.46, 0.88]) at follow-up, indicating acceptable internal consistency (Cronbach, 1951). The R package *ltm* (Rizopoulos, 2007) was used to calculate Cronbach's alpha with 10'000 permutations for the bootstrap 95%-CI.

*Inflammation measures*

Blood samples were collected after at least 3 hours of fasting. Inflammation markers were measured in plasma using enzyme-linked immunosorbent assays (ELISAs) and multiplex electrochemiluminescence systems. Specifically, plasma levels of CRP, leptin, and MIF (#DY289) were quantified using ELISAs from R&D systems (Minneapolis, USA). IL-6 was determined using multiplex electrochemiluminescence systems, specifically a SP-X™

imaging system from Quanterix (Billerica, USA). The chemokine CCL3 was determined using a MESO QuickPlex SQ 120MM apparatus (MesoScale Discovery, Rockville, Maryland, USA).

At baseline, three participants had missing blood samples, and one participant had a technical outlier in the MIF measure. These values were replaced with data collected on the day of surgery (4 weeks after scanning). For two participants, follow-up inflammation markers were not available, so scores were imputed using the sex-specific sample mean. To ensure that inflammation levels at baseline were due to chronic rather than acute inflammation, participants with extreme but transient values were identified as outliers. Specifically, participants with inflammation levels exceeding 3 standard deviations above (for CCL3, CRP, IL-6, IL-8, Leptin) or below (for MIF) the mean (equivalent to the top 0.15% in a normal distribution) at baseline, who also showed extreme changes in inflammation levels between baseline and surgery (exceeding 3 standard deviations), were considered outliers. One participant exhibited a severe but transient elevation in CRP, likely attributable to an acute infection (Ishii et al., 2012), while two participants displayed significantly heightened but transient IL-6 levels. These participants were excluded from further analyses (**Figure S1**).

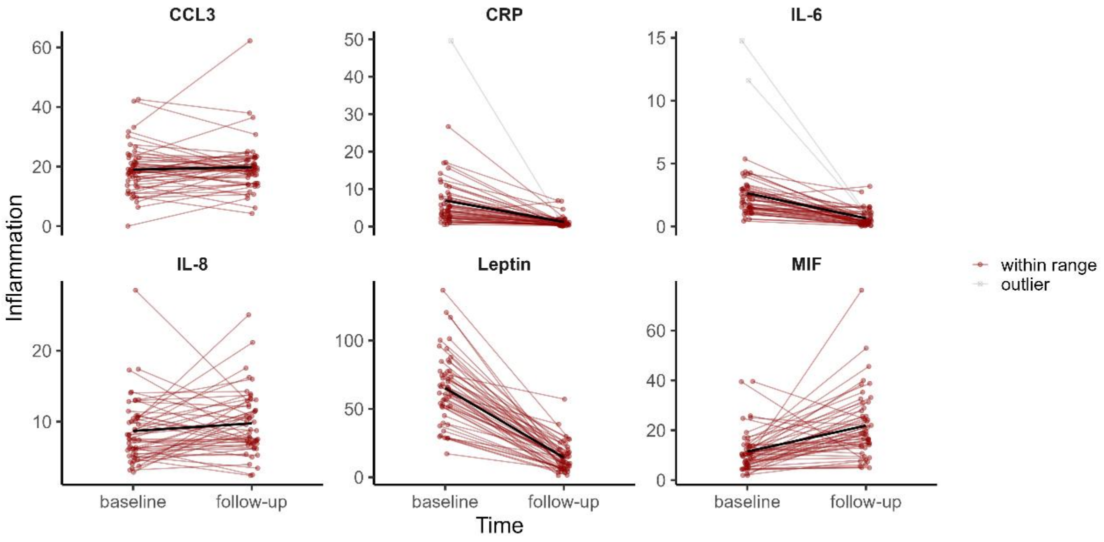

**Figure S1. Stability in inflammation levels over time.** The thin lines depict the individual inflammation trajectories (n=50). The bold black line represents the regression trend across all participants. At baseline, one participant exhibited a severe but transient elevation in CRP levels (> 49.5 mg/L) and two participants displayed strongly heightened but transient IL-6 levels. These three participants were excluded from further analyses.

##### MRI data acquisition

Imaging data were acquired using a 3.0 Tesla whole-body magnetic resonance imaging system (Magnetom Skyra, Siemens, HealthcareSector, Erlangen, Germany) equipped with a 32-channel head coil. Whole-brain structural images were acquired using a T1-weighted 3D magnetization-prepared rapid gradient-echo sequence (TR/TI/TE 2300/1100/3.03 ms; 8° flip angle; voxel size: 1.0×1.0×1.0 mm), and functional images were acquired using a T2*-weighted multiband, multi-echo planar imaging sequence to measure BOLD contrast during the Stroop task (TR/TE 1500/12.4, 34.3, 56.2 ms; 75° flip angle; voxel size: 2.5×2.5×2.5 mm; field of view 210 mm; 51 transversal slices in interleaved order).

##### MRI data preprocessing

Preprocessing of MRI data was performed using fMRIPrep 23.0.2 (Esteban et al., 2019, 2022) (RRID:SCR_016216), based on Nipype 1.8.4 (Gorgolewski et al., 2011) (RRID:SCR_002502). In brief, T1-weighted (T1w) images were corrected for intensity non-uniformity (Tustison et al., 2010), and used as T1w-reference throughout the workflow. The T1w-reference was then skull-stripped with OASIS30ANTs as target template, and segmented into tissue types (FSL 6.0.5.1:57b01774, RRID:SCR_002823). An anatomical T1w-reference map was computed after registration of the 2 T1w images per participant (after intensity non-uniformity-correction) using `mri_robust_template` (FreeSurfer 7.3.2, Reuter, Rosas, and Fischl 2010). Brain surfaces were reconstructed using `recon-all` (FreeSurfer 7.2.0, RRID:SCR_001847), and the brain mask estimated previously was refined to reconcile ANTs-derived and FreeSurfer-derived cortical gray-matter segmentations of Mindboggle (Klein et al., 2017) (RRID:SCR_002438). A FLAIR image was used to improve pial surface refinement. Images were spatially normalized to the *MNI152NLin6Asym* template (Evans et al., 2012) (RRID:SCR_002823) and resampled to 2 mm isotropic voxels.

Functional images were preprocessed by first generating a reference volume and its skull-stripped version from the shortest echo. Head-motion parameters (transformation matrices, and six corresponding rotation and translation parameters) were estimated with respect to the reference volume before any spatiotemporal filtering using `mcflirt` (FSL 6.0.5.1:57b01774) (Jenkinson et al., 2002). Functional images were slice-time corrected to 0.696s (0.5 of slice acquisition range 0s-1.39s) using `3dTshift` from AFNI (Cox & Hyde, 1997) (RRID:SCR_005927). The functional images were then resampled onto their original, native space by applying the transforms to correct for head-motion. A T2* map was estimated from these preprocessed EPI echoes, by voxel-wise fitting the maximal number of echoes with reliable signal in that voxel to a monoexponential signal decay model with nonlinear regression, and then used to optimally combine the preprocessed echoes (Posse et al., 1999). The functional reference volume was subsequently co-registered to the T1w-reference using `bbregister` (FreeSurfer) which implements boundary-based registration (Greve & Fischl, 2009). Co-registration was configured with six degrees of freedom. First, a reference volume and its skull-stripped version were generated using a custom methodology of fMRIPrep. Based on the preprocessed functional images, the following confounding time-series were extracted from the cerebrospinal fluid and white matter masks and written to a confounds file: framewise displacement (FD), DVARS and three region-wise global signals. FD was computed using the formulation following Jenkinson (relative root mean square displacement between affines, Jenkinson et al. (2002)). FD and DVARS were calculated for each functional run, both using their implementations in Nipype. The three global signals were extracted within the CSF, the WM, and the whole-brain masks. Additionally, automatic removal of motion artifacts using independent component analysis (ICA-AROMA) (Pruim et al., 2015) was performed and the noise-regressors written to the confounds file. The BOLD time-series were resampled into standard *MNI152NLin6Asym* space. Image quality was evaluated using visual reports from fMRIPrep.

**Analysis of fMRI activation**

Further processing was performed using SPM12 (version 7771; RRID:SCR_007037). The first 30 volumes of the functional images, acquired before the experiment started to facilitate echo combination, were removed to allow for signal stabilization, and the remaining time series were spatially smoothed with a 6mm full-width-half-maximum (FWHM) Gaussian kernel. First-level analysis was performed using a general linear model for each subject, containing separate regressors for the correct *congruent* and the correct *incongruent* trials, plus two regressors for missed and incorrect trials. All correct trials were modeled with a stick function aligned to the onset of each stimulus, convolved with the canonical hemodynamic response function. To account for head motion, confound time-series from cerebrospinal fluid and white matter masks, and automatic removal of motion artifacts using independent component analysis (ICA-AROMA) (Pruim et al., 2015), were included as nuisance regressors. Low-frequency drifts were removed with a high-pass filter at 128s, and serial correlations in the time series were accounted for using a first-degree autoregressive model.

**Inflammation-sensitive mask**

To investigate the potential link between changes in systemic inflammation and changes in neural activation, we focused the second-level analysis on regions sensitive to inflammatory markers. Specifically, we utilized a map of empirically-derived brain regions sensitive to systemic inflammation, accessible on NeuroVault (https://identifiers.org/neurovault.collection:3234). A map with a stringent extent-based threshold (voxel-wise α = 0.001) was selected and binarized. Subsequently, we employed it to select all parcels from the Automated Anatomical Labeling atlas 3v1 (Rolls et al., 2020), that contained at least one significant voxel. This selection process allowed for the creation of an inflammation-sensitive brain mask, while ensuring meaningful and comprehensive brain parcels. See **Table S1** for a full list of all included parcels. The binarized map is available on NeuroVault (RRID:SCR_003806; https://identifiers.org/neurovault.image:868591). Further methodological details can be found in Kraynak et al. (2018).

**Table S1**. List of inflammation-sensitive brain regions included in the mask

| # | Region label | Description | # | Region label | Description |
|---|---|---|---|---|---|
| 3 | Frontal_Sup_2_L | Superior frontal gyrus, dorsolateral | 75 | Caudate_L | Caudate nucleus |
| 5 | Frontal_Mid_2_L | Middle frontal gyrus | 77 | Putamen_L | Lenticular nucleus, Putamen |
| 8 | Frontal_Inf_Oper_R | Inferior frontal gyrus, opercular part | 78 | Putamen_R | Lenticular nucleus, Putamen |
| 10 | Frontal_Inf_Tri_R | Inferior frontal gyrus, triangular part | 79 | Pallidum_L | Lenticular nucleus, Pallidum |
| 12 | Frontal_Inf_Orb_2_R | IFG pars orbitalis, | 83 | Heschl_L | Heschl’s gyrus |

| | | | | | |
|---|---|---|---|---|---|
| 14 | Rolandic_Oper_R | Rolandic operculum | 85 | Temporal_Sup_L | Superior temporal gyrus |
| 15 | Supp_Motor_Area_L | Supplementary motor area | 86 | Temporal_Sup_R | Superior temporal gyrus |
| 17 | Olfactory_L | Olfactory cortex | 88 | Temporal_Pole_Sup_R | Temporal pole: superior temporal gyrus |
| 18 | Olfactory_R | Olfactory cortex | 90 | Temporal_Mid_R | Middle temporal gyrus |
| 19 | Frontal_Sup_Medial_L | Superior frontal gyrus, medial | 92 | Temporal_Pole_Mid_R | Temporal pole: middle temporal gyrus |
| 20 | Frontal_Sup_Medial_R | Superior frontal gyrus, medial | 93 | Temporal_Inf_L | Inferior temporal gyrus |
| 21 | Frontal_Med_Orb_L | Superior frontal gyrus, medial orbital | 94 | Temporal_Inf_R | Inferior temporal gyrus |
| 23 | Rectus_L | Gyrus rectus | 96 | Cerebellum_Crus1_R | Crus I of cerebellar hemisphere |
| 25 | OFCmed_L | Medial orbital gyrus | 101 | Cerebellum_4_5_L | Lobule IV, V of cerebellar hemisphere |
| 29 | OFCpost_L | Posterior orbital gyrus | 102 | Cerebellum_4_5_R | Lobule IV, V of cerebellar hemisphere |
| 30 | OFCpost_R | Posterior orbital gyrus | 104 | Cerebellum_6_R | Lobule VI of cerebellar hemisphere |
| 33 | Insula_L | Insula | 125 | Thal_VA_L | Ventral anterior |
| 34 | Insula_R | Insula | 127 | Thal_VL_L | Ventral lateral |
| 37 | Cingulate_Mid_L | Middle cingulate & paracingulate gyri | 129 | Thal_VPL_L | Ventral posterolateral |
| 38 | Cingulate_Mid_R | Middle cingulate & paracingulate gyri | 131 | Thal_IL_L | Intralaminar |
| 41 | Hippocampus_L | Hippocampus | 139 | Thal_LGN_L | Lateral geniculate |
| 42 | Hippocampus_R | Hippocampus | 141 | Thal_MGN_L | Medial Geniculate |
| 43 | ParaHippocampal_L | Parahippocampal gyrus | 143 | Thal_PuI_L | Pulvinar anterior |
| 44 | ParaHippocampal_R | Parahippocampal gyrus | 145 | Thal_PuM_L | Pulvinar medial |
| 45 | Amygdala_L | Amygdala | 151 | ACC_sub_L | Anterior cingulate cortex, subgenual |
| 46 | Amygdala_R | Amygdala | 153 | ACC_pre_L | Anterior cingulate cortex, pregenual |
| 51 | Lingual_L | Lingual gyrus | 155 | ACC_sup_L | Anterior cingulate cortex, supracallosal |
| 59 | Fusiform_L | Fusiform gyrus | 157 | N_Acc_L | Nucleus accumbens |
| 60 | Fusiform_R | Fusiform gyrus | 161 | SN_pc_L | Substantia nigra, pars compacta |
| 71 | Precuneus_L | Precuneus | 163 | SN_pr_L | Substantia nigra, pars reticulata |

*Note*. Labels from the Automated Anatomical Labeling atlas 3v1 (Rolls et al., 2020).

**Analysis of fMRI connectivity**

To investigate task-modulated connectivity, i.e., changes in functional association strength covarying with response conflict processing, generalized psychophysiological interaction (gPPI) analyses were conducted (McLaren et al., 2012). The five clusters with the strongest activation differences between conditions at baseline were used as seed regions. For each participant, multiple regression models were calculated between the seed regions and every gray-matter voxel of the brain, including the following regressors: i) the two task conditions convolved with the canonical hemodynamic response function, ii) the eigenvariate of the time series from the seed region, and iii) the psycho-physiological interaction terms specified as product of i) and ii). For each participant, one [follow-up > baseline] contrast image [(incongruent$_{follow\text{-}up}$ - congruent$_{follow\text{-}up}$) - (incongruent$_{baseline}$ - congruent$_{baseline}$)] was generated, containing voxel-wise estimates of changes in functional connectivity modulation during response conflict processing.

### Supplemental Results

**Inflammation results**

*Intercorrelations of inflammation markers*

To assess the intercorrelations of the change in inflammation markers, pairwise partial correlations were calculated between change scores of all measures (**Figure S3**).

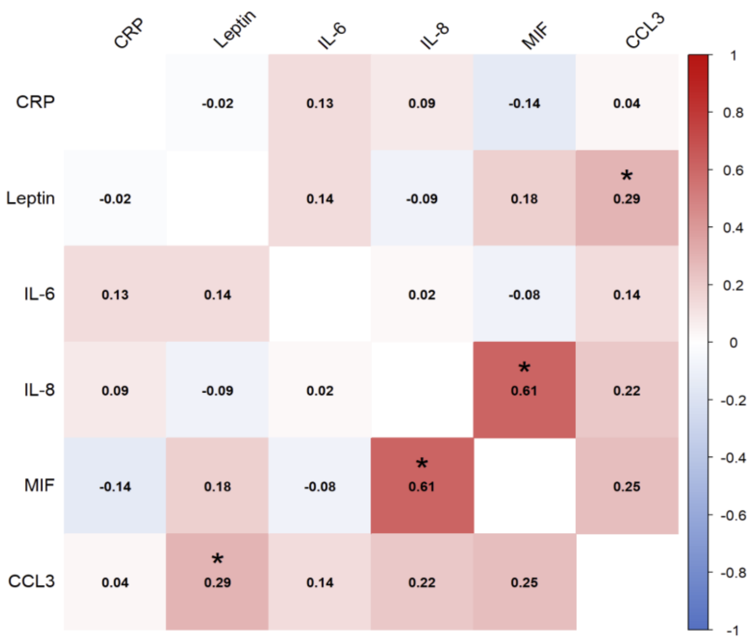

**Figure S2. Intercorrelations of change in inflammation markers.** The correlation matrix displays partial Spearman's *rho* for all pairwise correlations, controlled for age and sex. n=47. ∗ $p < 0.05$.

**Neuroimaging results**

*Brain activation during response conflict*

**Table S2. Stroop effect at baseline.** Summary of brain regions exhibiting main task effects with a cluster size ≥ 10 voxels. Coordinates of the peak of a cluster (X, Y, Z) are given in MNI space.

| **Region** | **Side (L/R)** | | **X,Y,Z** | | ***T*** | **Size (# voxels)** | ***p*FWE** |
|---|---|---|---|---|---|---|---|
| MFG | L | -34 | 4 | 64 | 7.30 | 509 | <.0001 |
| SMA | L | -6 | 14 | 54 | 6.99 | 1222 | <.0001 |
| Insula | R | 30 | 22 | -10 | 6.90 | 783 | <.0001 |
| MFG, inferior | L | -48 | 12 | 36 | 6.90 | 170 | <.0001 |
| Caudate nucleus | R | 14 | 8 | 6 | 6.80 | 236 | <.0001 |
| Insula | L | -40 | 16 | -2 | 6.38 | 809 | <.0001 |
| Occipital inferior gyrus | L | -46 | -66 | -14 | 6.19 | 287 | <.0001 |
| IFG, opercular part | R | 54 | 20 | 32 | 6.18 | 374 | <.0001 |
| Thalamus | L | -12 | -18 | 16 | 5.49 | 79 | .033 |
| Middle temporal gyrus | L | -64 | -32 | 4 | 4.47 | 211 | <.0001 |
| Occipital inferior gyrus | R | 38 | -86 | -2 | 4.81 | 143 | .002 |

*Note*. Results were anatomically labelled by reference to the Automated Anatomical Labelling atlas 3v1 (Rolls et al., 2020). IFG = Inferior frontal gyrus, MFG= Middle frontal gyrus, SMA= Supplemental Motor Area. L=left, R=right.

*Relationship between brain activation and inflammation*

**Table S3. Relationship between brain activation and inflammation.** Summary of fixed effects from linear mixed-effect models.

| Region | Parameter | Coefficient | SE | CI_low | CI_high | *t*(86) | *p* | *q* [1] |
|---|---|---|---|---|---|---|---|---|
| Caudate | CRP | 0.033 | 0.042 | -0.049 | 0.116 | 0.806 | 0.423 | 1.000 |
| Caudate | time x CRP | -0.151 | 0.156 | -0.461 | 0.160 | -0.965 | 0.337 | 1.000 |
| Caudate | IL-6 | -0.071 | 0.054 | -0.177 | 0.035 | -1.327 | 0.188 | 0.564 |
| Caudate | time x IL-6 | 0.025 | 0.096 | -0.165 | 0.216 | 0.265 | 0.792 | 1.000 |
| Caudate | Leptin | -0.053 | 0.064 | -0.180 | 0.074 | -0.830 | 0.409 | 0.919 |
| Caudate | time x Leptin | 0.090 | 0.151 | -0.210 | 0.390 | 0.598 | 0.551 | 1.000 |
| Caudate | MIF | -0.039 | 0.071 | -0.180 | 0.103 | -0.543 | 0.588 | 1.000 |
| Caudate | time x MIF | 0.060 | 0.082 | -0.103 | 0.224 | 0.731 | 0.466 | 0.840 |
| Insula L | CRP | 0.011 | 0.034 | -0.057 | 0.079 | 0.316 | 0.753 | 1.000 |
| Insula L | time x CRP | 0.057 | 0.131 | -0.202 | 0.317 | 0.438 | 0.663 | 1.000 |
| Insula L | IL-6 | -0.070 | 0.044 | -0.157 | 0.018 | -1.577 | 0.118 | 0.564 |
| Insula L | time x IL-6 | 0.054 | 0.080 | -0.104 | 0.212 | 0.682 | 0.497 | 1.000 |
| Insula L | Leptin | -0.049 | 0.053 | -0.154 | 0.056 | -0.926 | 0.357 | 0.919 |
| Insula L | time x Leptin | -0.012 | 0.126 | -0.263 | 0.239 | -0.094 | 0.925 | 1.000 |
| Insula L | MIF | -0.041 | 0.058 | -0.156 | 0.075 | -0.700 | 0.486 | 1.000 |
| Insula L | time x MIF | 0.074 | 0.068 | -0.061 | 0.209 | 1.094 | 0.277 | 0.681 |
| Insula R | CRP | 0.037 | 0.037 | -0.037 | 0.110 | 0.988 | 0.326 | 1.000 |
| Insula R | time x CRP | -0.096 | 0.139 | -0.373 | 0.182 | -0.686 | 0.495 | 1.000 |
| Insula R | IL-6 | -0.052 | 0.048 | -0.147 | 0.042 | -1.097 | 0.276 | 0.620 |
| Insula R | time x IL-6 | -0.078 | 0.085 | -0.248 | 0.091 | -0.917 | 0.362 | 1.000 |
| Insula R | Leptin | -0.088 | 0.057 | -0.201 | 0.024 | -1.558 | 0.123 | 0.554 |
| Insula R | time x Leptin | 0.063 | 0.134 | -0.203 | 0.329 | 0.472 | 0.638 | 1.000 |
| Insula R | MIF | -0.046 | 0.063 | -0.171 | 0.079 | -0.727 | 0.469 | 1.000 |
| Insula R | time x MIF | 0.097 | 0.073 | -0.048 | 0.243 | 1.330 | 0.187 | 0.681 |
| MFG | CRP | -0.023 | 0.036 | -0.095 | 0.049 | -0.641 | 0.524 | 1.000 |
| MFG | time x CRP | 0.002 | 0.139 | -0.275 | 0.279 | 0.013 | 0.990 | 1.000 |
| MFG | IL-6 | -0.064 | 0.047 | -0.157 | 0.029 | -1.375 | 0.173 | 0.564 |
| MFG | time x IL-6 | 0.119 | 0.084 | -0.049 | 0.287 | 1.407 | 0.163 | 1.000 |
| MFG | Leptin | -0.009 | 0.056 | -0.120 | 0.102 | -0.160 | 0.874 | 1.000 |
| MFG | time x Leptin | -0.014 | 0.134 | -0.280 | 0.252 | -0.105 | 0.917 | 1.000 |
| MFG | MIF | -0.067 | 0.061 | -0.188 | 0.054 | -1.107 | 0.271 | 1.000 |
| MFG | time x MIF | 0.107 | 0.071 | -0.034 | 0.249 | 1.505 | 0.136 | 0.681 |
| SMA | CRP | -0.005 | 0.034 | -0.073 | 0.063 | -0.142 | 0.888 | 1.000 |
| SMA | time x CRP | -0.054 | 0.130 | -0.312 | 0.204 | -0.417 | 0.678 | 1.000 |
| SMA | IL-6 | -0.032 | 0.044 | -0.119 | 0.056 | -0.713 | 0.478 | 0.860 |
| SMA | time x IL-6 | 0.048 | 0.079 | -0.110 | 0.206 | 0.607 | 0.545 | 1.000 |
| SMA | Leptin | -0.093 | 0.052 | -0.196 | 0.010 | -1.790 | 0.077 | 0.554 |

| Region | Parameter | Coefficient | SE | CI_low | CI_high | *t*(86) | *p* | *q* [1] |
|---|---|---|---|---|---|---|---|---|
| SMA | time x Leptin | 0.110 | 0.123 | -0.134 | 0.354 | 0.894 | 0.374 | 1.000 |
| SMA | MIF | -0.035 | 0.058 | -0.150 | 0.079 | -0.612 | 0.542 | 1.000 |
| SMA | time x MIF | 0.070 | 0.067 | -0.064 | 0.203 | 1.037 | 0.303 | 0.681 |

*Note.* Parameters were z-standardized before analyses. [1] FDR-corrected for five regions of interest and four inflammation markers.

*Relationship between brain activation and performance*

**Table S4. Relationship between brain activation and performance.** Summary of effects of interest from linear mixed-effect models.

| Region | Parameter | Coefficient | SE | CI_low | CI_high | *t*(86) | *p* | *q* [1] |
|---|---|---|---|---|---|---|---|---|
| Caudate | accuracy Stroop | -0.070 | 0.041 | -0.152 | 0.012 | -1.705 | 0.092 | 0.459 |
| Caudate | time x accuracy Stroop | -0.006 | 0.068 | -0.142 | 0.130 | -0.089 | 0.929 | 0.929 |
| Caudate | response time Stroop | 0.089 | 0.047 | -0.005 | 0.183 | 1.887 | 0.063 | 0.313 |
| Caudate | time x rt Stroop | -0.057 | 0.062 | -0.181 | 0.066 | -0.923 | 0.359 | 0.839 |
| Insula L | accuracy Stroop | -0.013 | 0.034 | -0.080 | 0.054 | -0.376 | 0.708 | 0.708 |
| Insula L | time x accuracy Stroop | -0.088 | 0.057 | -0.200 | 0.024 | -1.555 | 0.124 | 0.614 |
| Insula L | response time Stroop | 0.002 | 0.040 | -0.077 | 0.080 | 0.037 | 0.970 | 0.970 |
| Insula L | time x rt Stroop | 0.011 | 0.054 | -0.096 | 0.118 | 0.204 | 0.839 | 0.839 |
| Insula R | accuracy Stroop | -0.030 | 0.037 | -0.103 | 0.043 | -0.824 | 0.412 | 0.688 |
| Insula R | time x accuracy Stroop | -0.059 | 0.061 | -0.181 | 0.063 | -0.965 | 0.338 | 0.614 |
| Insula R | response time Stroop | 0.049 | 0.042 | -0.036 | 0.133 | 1.148 | 0.254 | 0.452 |
| Insula R | time x rt Stroop | -0.015 | 0.057 | -0.128 | 0.098 | -0.260 | 0.795 | 0.839 |
| MFG | accuracy Stroop | 0.034 | 0.036 | -0.038 | 0.105 | 0.933 | 0.353 | 0.688 |
| MFG | time x accuracy Stroop | -0.046 | 0.060 | -0.166 | 0.074 | -0.768 | 0.445 | 0.614 |
| MFG | response time Stroop | 0.023 | 0.041 | -0.058 | 0.104 | 0.576 | 0.566 | 0.708 |
| MFG | time x rt Stroop | 0.043 | 0.057 | -0.070 | 0.156 | 0.753 | 0.453 | 0.839 |
| SMA | accuracy Stroop | 0.014 | 0.034 | -0.053 | 0.082 | 0.422 | 0.674 | 0.708 |
| SMA | time x accuracy Stroop | -0.039 | 0.057 | -0.152 | 0.074 | -0.691 | 0.491 | 0.614 |
| SMA | response time Stroop | 0.043 | 0.039 | -0.034 | 0.119 | 1.107 | 0.272 | 0.452 |
| SMA | time x rt Stroop | 0.023 | 0.053 | -0.082 | 0.128 | 0.429 | 0.669 | 0.839 |

*Note.* Parameters were z-standardized before analyses. [1] FDR-corrected for five regions of interest.

*Task connectivity during response conflict*

**Table S5**. Summary of inflammation-specific functional connectivity analyses. Coordinates of the peak of a cluster (X, Y, Z) are given in MNI space.

| Cov | Seed | Region | Side (L/R) | X,Y,Z | | | *T* | Size (# voxels) | pFWE |
|---|---|---|---|---|---|---|---|---|---|
| MIF | Caudate R | SFGmed | L | 10 | 50 | 22 | 4.97 | 121 | <.005 |
| Leptin | Insula L | SFGmed | L | -6 | 40 | 10 | 4.59 | 138 | <.005 |

*Note*. Results were anatomically labelled by reference to the Automated Anatomical Labelling atlas 3v1 (Rolls et al., 2020). All models were controlled for age and sex. The MIF-related cluster in the left superior frontal gyrus was spanning both hemispheres, covering also parts of the right anterior cingulate gyrus. Cov=covariate of interest, SFG = Superior frontal gyrus, SFGmed = Superior frontal gyrus, medial, SMA= Supplemental Motor Area. L=left, R=right.

*Relationship between task connectivity, inflammation, and performance*

**Table S6**. **Relationship between inflammation-specific connectivity changes and performance.** Summary of effects of interest from linear mixed-effect models.

| Region | Parameter | Coefficient | SE | CI_low | CI_high | *t*(86) | *p* | *q* [1] |
|---|---|---|---|---|---|---|---|---|
| Leptin: SFGmed | accuracy Stroop | -0.032 | 0.156 | -0.342 | 0.278 | -0.205 | 0.838 | 1.000 |
| Leptin: SFGmed | time x accuracy Stroop | -0.269 | 0.258 | -0.782 | 0.245 | -1.039 | 0.301 | 0.603 |
| Leptin: SFGmed | response time Stroop | -0.303 | 0.178 | -0.657 | 0.050 | -1.705 | 0.092 | 0.184 |
| Leptin: SFGmed | time x response time Stroop | 0.439 | 0.239 | -0.036 | 0.913 | 1.837 | 0.070 | 0.139 |
| MIF: SFGmed | accuracy Stroop | 0.373 | 0.155 | 0.066 | 0.681 | 2.416 | 0.018 | 0.036 |
| MIF: SFGmed | time x accuracy Stroop | -0.702 | 0.257 | -1.212 | -0.191 | -2.732 | 0.008 | 0.015 |
| MIF: SFGmed | response time Stroop | -0.212 | 0.184 | -0.579 | 0.155 | -1.148 | 0.254 | 0.508 |
| MIF: SFGmed | time x response time Stroop | 0.329 | 0.250 | -0.168 | 0.826 | 1.318 | 0.191 | 0.382 |

*Note.* Parameters were z-standardized before analyses. [1] FDR-corrected for two regions of interest. SFGmed= Superior frontal gyrus, medial.
